# Cost-utility of aripiprazole once-monthly versus paliperidone palmitate once-monthly injectable for schizophrenia in China

**DOI:** 10.1101/2024.12.30.24319781

**Authors:** Yiping An, Jinxi Ding, Wei Li, Gang Fang, Zhipeng Pi, Yumeng Zhang

## Abstract

**Objectives:** From the perspective of Chinese healthcare system, this study compared the cost-utility of aripiprazole once-monthly (AOM) and paliperidone palmitate once-monthly injectable for the treatment of adult Chinese patients with schizophrenia.

**Methods:** A 5-state Markov model was constructed to assess the cost-utility of 10 years of treatment with long-acting injections (LAI) for schizophrenia. The long-term costs and quality-adjusted life years (QALYs) were estimated. The outcome was the incremental cost-effectiveness ratio (ICER). The annual discount rate was set at 5 %. Willingness-to-pay (WTP) threshold of 1x GDP was used to judge the economics of intervention.

**Results:** In the base-case, the incremental costs of AOM relative to PP1M after 10 years of treatment were US$1,926.373 with an incremental QALYs of 0.306. The ICER for AOM was US$6,285.303/QALY, which was lower than the WTP threshold of US$12,538.306/QALY. The one-way sensitivity analysis and probability sensitivity analysis verified the robustness of the base-case. Scenario analysis showed that from 10 to 30 years, as the horizon increased, the ICER became smaller, and all were below the WTP.

**Conclusions:** This analysis showed that compared with PP1M, AOM represents a long-term cost-utility advantages for schizophrenia treatment in China.

## Introduction

The World Health Organization (WHO) reports that schizophrenia affects approximately 24 million people globally.(1) Meanwhile, ‘The National Information System for Psychosis’ indicates that the condition impacts about 4.6 million individuals in China.(2) An epidemiological study of covering 195 countries and territories said that China has one of the highest prevalence rates in the world. (3) Schizophrenia often develops in young adults in their twenties, severely affecting personal functions and exacerbating the loss of productivity in an ageing society.

Schizophrenia has a high disease burden. It is characterized by poor treatment adherence and easy to relapse. The three-year compliance rate was only 40.87 %, and the three-year relapse rate was highly 61.74 %.(4) A disease history of multiple relapse is associated with a poorer prognosis for patients, indicating that patients require a longer treatment period to achieve the same efficacy as before the relapses, (5) this exacerbates the burden on patients.

The latest Chinese clinical guideline has recommended that atypical antipsychotic LAI should be applied as first-line strategy for patients with acute and maintenance schizophrenia. Patients who received LAI early in the course of their illness had significantly lower rate of hospitalization and treatment interruption than those who received oral antipsychotics.(6)

AOM is the newest third-generation atypical LAI launched in China in 2023, and the world’s first partial agonist of dopamine D2 and 5HT-1A receptors, compared to the second-generation paliperidone and risperidone long-acting injection, both of which are full 5HT-2A and D2 receptor antagonists. In terms of mechanism of action, partial agonists bind to the D2 receptor more strongly than full antagonists. This makes them more effective than full antagonists at ameliorating positive symptoms and the risk of high prolactin levels. This also improve negative, cognitive, and affective symptoms more favorably than full antagonists, which primarily ameliorate positive symptoms.(7) AOM achieves a balance of agonistic and antagonistic effects, making it a dopamine system stabilizer. A positively controlled (AOM vs oral aripiprazole), randomized, double-blind phase III marketed clinical trial(8) (NCT03172871) demonstrated the efficacy and safety of AOM in Chinese acute schizophrenia patients.

PP1M was launched in China in 2011 and has been included in the China National Reimbursement Drug List (NRDL) in 2020, and has a post-marketing phase IV single-arm, open-label trial in China, also for acute schizophrenia.(9) PP1M is highly similar to AOM. They have the same indication, route of administration, frequency of dosing, and sequence of treatment. Furthermore, PP1M is also a standard therapeutic drug, recommended by domestic and international guidelines.(10–12) Therefore, this study aims to evaluate the cost-utility of AOM compared to PP1M in treating Chinese adult schizophrenia from the perspective of the Chinese healthcare system.

## Methods

### Study Design

Compared with the decision tree model, the Markov model demonstrates the disease course in schizophrenia more completely, which is consistent with the characteristics of chronic diseases and long-acting injectable drugs. We used Excel 2021 to build a 5 state-transition Markov model (Fig 1) to evaluate the cost-utility of AOM and PP1M over a 10-year horizon. Matching-adjusted indirect comparisons (MAIC) of the AOM Phase III Trial and the PP1M Phase IV Trial to obtain acute efficacy and safety data. Meta-analysis of other placebo-controlled randomized clinical trials of AOM and PP1M to obtain relapse rate, and refer to published literature for other transition probability related parameters. Only the direct medical costs were considered, all costs are based on the official exchange rate of US$1 = 7.127 yuan.(The People’s Bank of China on June 30, 2024)(13) The mapping method and published literature were used to obtain utility values. The outcome of this study was ICER, which represents the costs of a schizophrenia patient per additional healthy year of survival. The annual discount rate for costs and utilities was set at 5 %. The willingness-to-pay threshold was US$12,538.306 as recommended by the WHO. (Based on the GDP one per capita in China in 2023.)(14)

**Figure 1.**
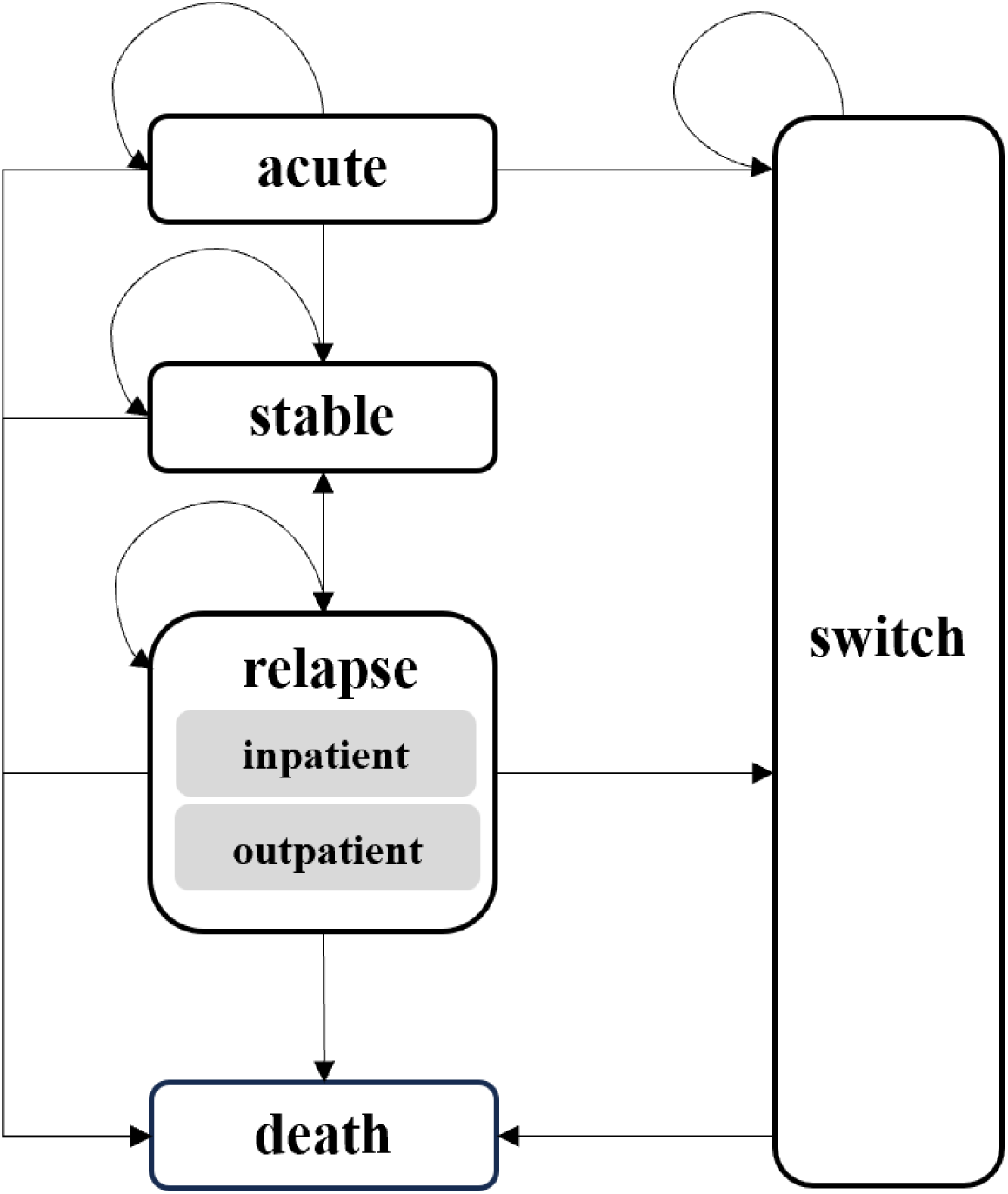
Markov model.

### Patient population

Patient characteristics were matched to the baseline patient cohort from our MAIC result. (See appendix 2 in the S1 File for details of MAIC.) Patients enter the model at age 31.5 years, with 55% of the cohort being male, BMI:23.2 kg/m^2^, Weight:64.4kg, Han Ethnicity:99%, PANSS total score:91.8, CGI-S total score:5.3, PSP total score:44.9.

### Model structure

The Markov model consists of five independent health states: (1) Acute, which is the initial state for all patients, is set to 12 weeks according to the timeframe of the AOM phase III trial study, and all patients are transferred out of this state after 12 weeks; (2) Stable; (3) Relapse, which includes both outpatient and inpatient; (4) Switch; (5) Death. The classification of health states is based on the ‘Diagnostic and Treatment Guidelines for Mental Disorders (2020 Edition)’,(15) which categorizes schizophrenia into four treatment states: acute, consolidation, maintenance, and chronic. (See appendix 1 in the SI File for details of the definitions of the health states and treatment modalities.)

The doses of the therapeutic drugs were derived from the product instructions. The choice of drugs for the switch state was informed by the consensus of clinical experts interviewed. (See appendix 4 in the SI File for details of clinical research.) They considered patients who transition from relapse to switch state to be more severely ill than those who transition from acute to switch state, and therefore these two states of patients are treated with different oral antipsychotics, with olanzapine being the last treatment option. A 1-month cycle was used based on the injection times specified in the instructions for AOM and PP1M. Half-cycle corrections were applied to each cycle to adjust the costs and QALYs to occur at the midpoint of the month.

### Clinical inputs and transition probabilities

The QUALIFY study was the only head-to-head clinical trial comparing AOM and PP1M that focused on quality of life.(16) Its primary outcomes, the Heinrichs-Carpenter Quality of Life Scale (QLS) scores and the Clinical Global Impression-Severity scale (CGI-S), were difficult to use for determining transition probabilities. Therefore, the study derived the transition probabilities through MAIC, meta-analysis, and other published literature, as shown in Table 1. All parameters were transformed and input into the model using Formula 1.(17)

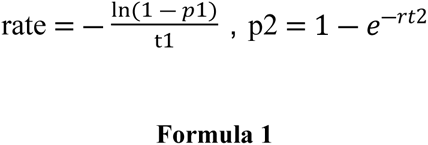

**Table 1.**
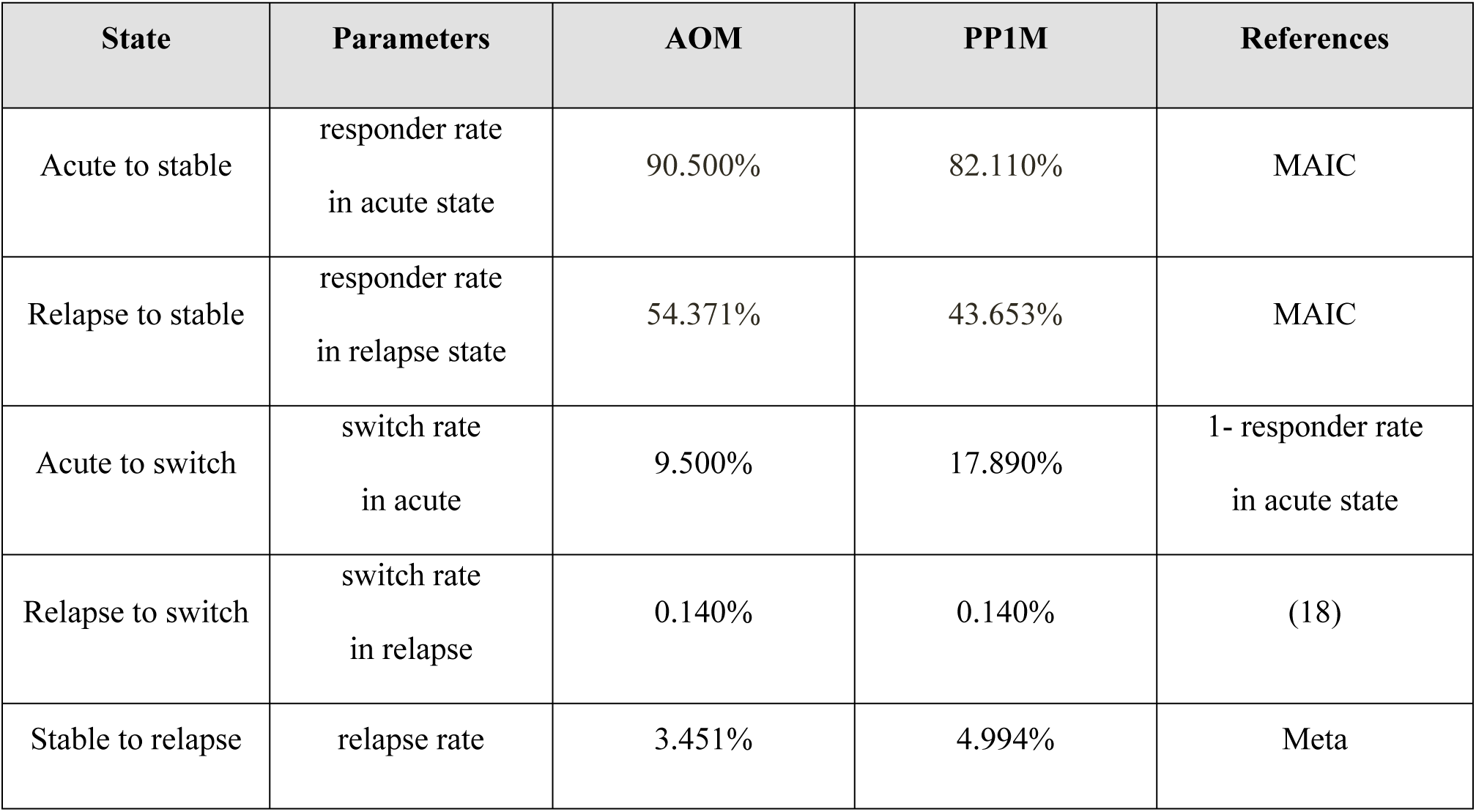
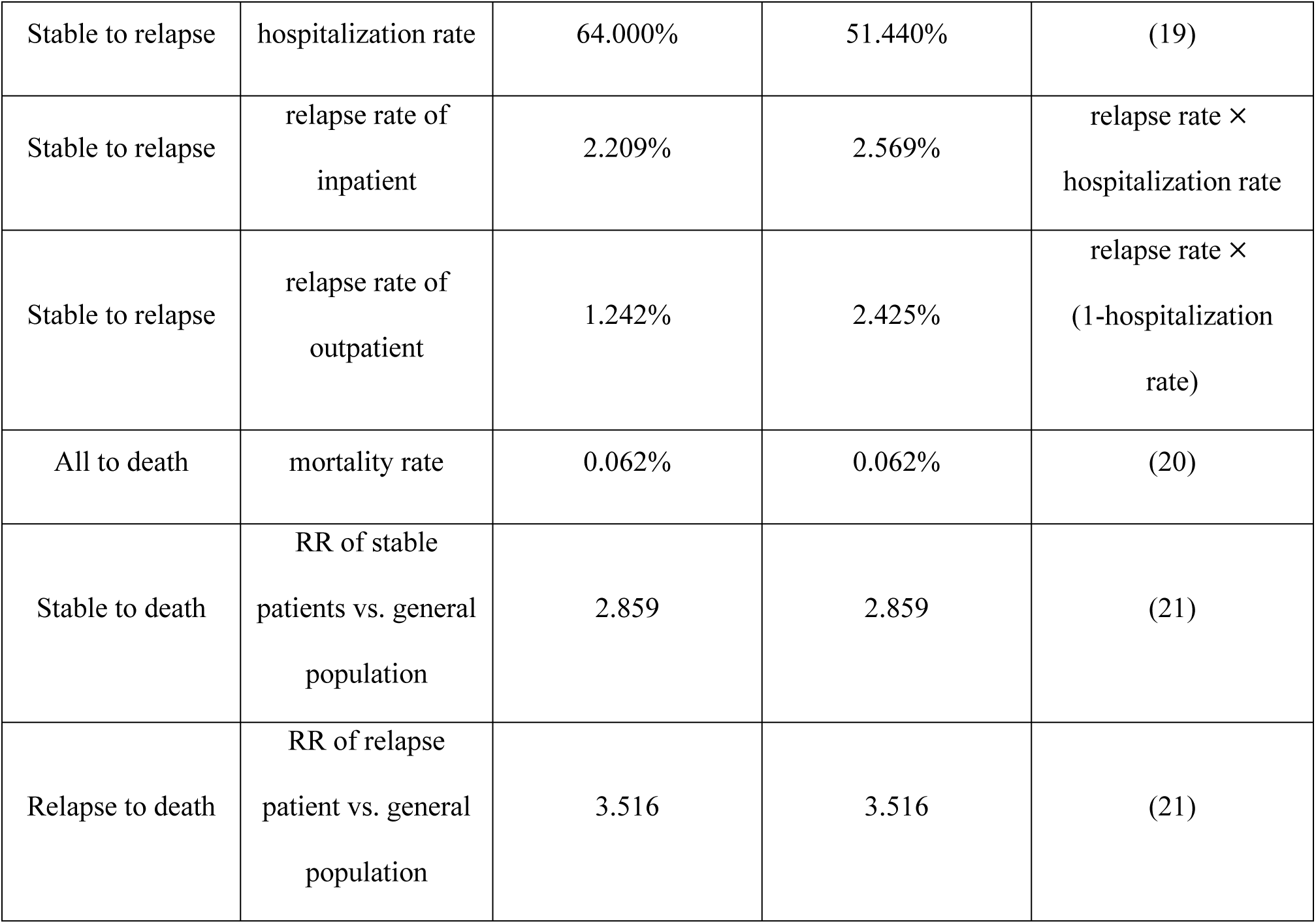
Transition probability.

rate: rate of occurrence of the event

p1: probability of an event occurring within the time frame of the study as reported in the original literature

t1: time frame for events reported in the original literature

p2: transition probability

t2: cycle period

The transition probability for the acute state was based on the responder rate, defined as ≥30 % reduction in the Positive and Negative Syndrome Scale (PANSS) total score.(8) This parameter was derived from the MAIC by adjusting individual patient data (IPD) from the AOM phase III trial and aggregated data (AgD) from the PP1M phase IV Trial. Both trials included patient populations with acute exacerbations of schizophrenia (baseline PANSS total score ≥70). The pre-adjusted responder rates were (AOM: 88.600 %, PP1M: 82.110 %), and the adjusted indirectly comparable responder rates were (AOM: 90.500 %, PP1M: 82.110 %). (See appendix 2 in the SI File for details of MAIC.)

Due to the short time horizon of the AOM phase III trial (12 weeks) and the PP1M phase IV trial (13 weeks), and the absence of efficacy and safety data in the relapse state, the study hypothesized that responder rate and adverse event rate in the relapse state would be equivalent to those in the acute state. Transition probability was adjusted according to Formula 1 to reflect this assumption. This assumption was endorsed by clinical experts and corroborated by published literature. Emsley et al(22) conducted a 7-year cohort study comparing treatment responder rate, remission rate, and total PANSS scores between patients using LAI for schizophrenia after a first episode and those after a relapse. The results showed no significant differences, leading to the conclusion that the responder rate to treatment in relapses were similar to those in the first episodes.

Since there are no published meta-analyses specifically addressing LAI, for schizophrenia that include relapse rate, we conducted a meta-analysis to derive relapse rate in the stable state. (See appendix 3 in the S1 File for details of meta.) This meta-analysis yielded a relative risk (RR) of relapse for AOM versus PP1M of 0.691 (95 % CI, 0.407-1.173), with relapse rate (AOM: 3.451 %, PP1M: 4.994 %). Calculate the transition probabilities for relapse inpatient and outpatient by multiplying the relapse rate and the hospitalization rate(AOM:64.000 %, PP1M 51.440 %).(19)

The mortality rate for the general population is derived from the latest China Statistical Yearbook and is 0.062%.(20) The transition probability of death state was calculated from the RR values of mortality in schizophrenia patients in the relapse state vs the general population(AOM:3.516, PP1M:3.092), and the RR values of mortality in schizophrenia patients in the stable phase vs the general population.(AOM:2.859, PP1M:2.622).(21)

In addition to the transition probability, the clinical inputs include the rates of adverse events as shown in Table 2. To simplify the model calculations, only three adverse events of particular interest from the AOM phase III trial were considered, including weight gain (defined as weight increased by≥7 %), prolactin-related adverse events (included decreased or increased blood prolactin levels, hyperprolactinemia), and extrapyramidal symptoms (EPS)-related adverse events (included dystonia, Parkinson disease, akathisia, muscle rigidity, etc). The adverse event rates for the acute and relapse states were derived from the MAIC. The adverse event rates for the stable and switch states were derived from the QUALIFY study.(16) This reported the aforementioned three adverse events in the AOM versus PP1M trial with a follow-up period of up to 32 weeks, which compensated for the short time frame in observing adverse event rates in the AOM phase III trial and the PP1M single-arm trial.

**Table 2.**
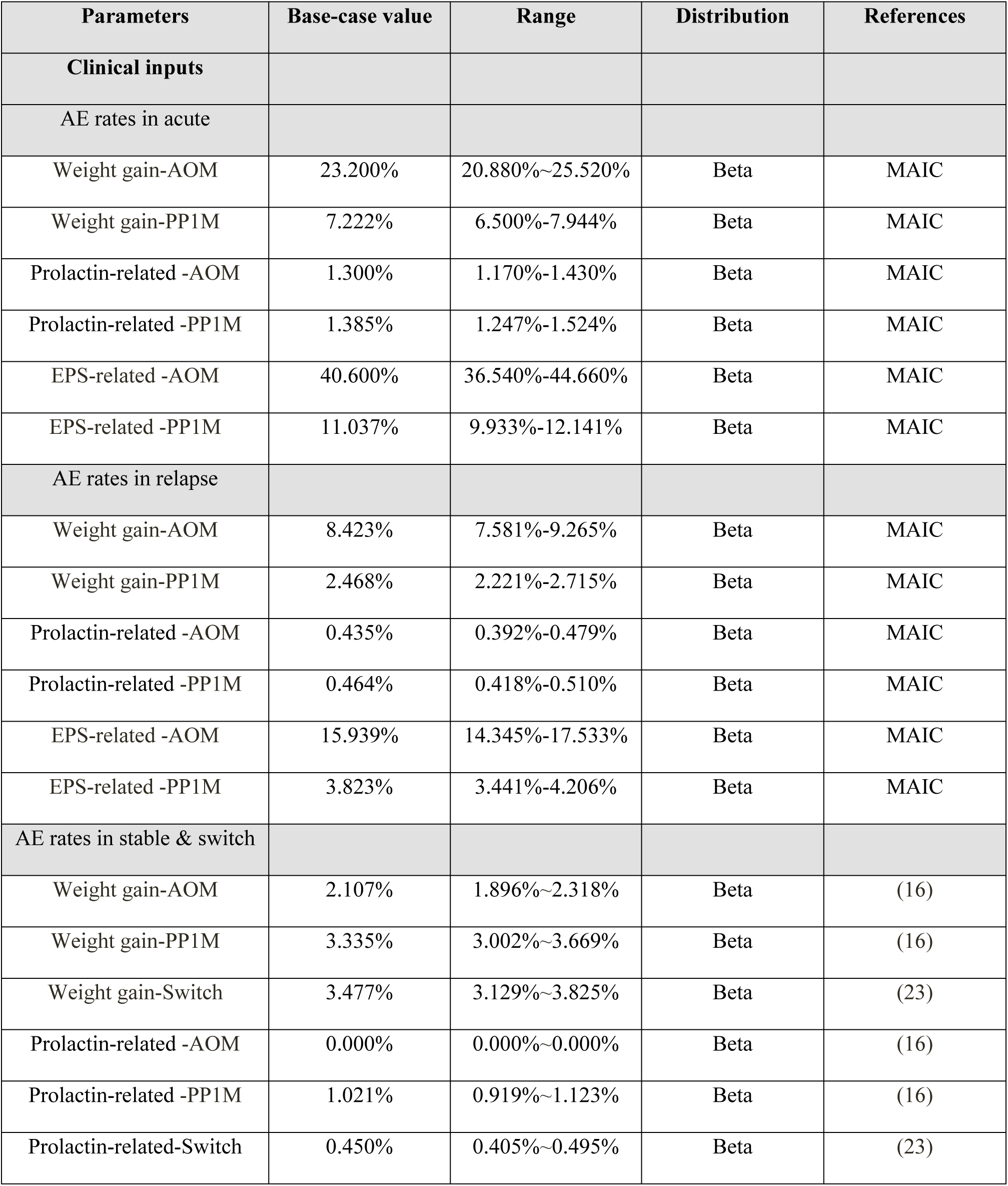
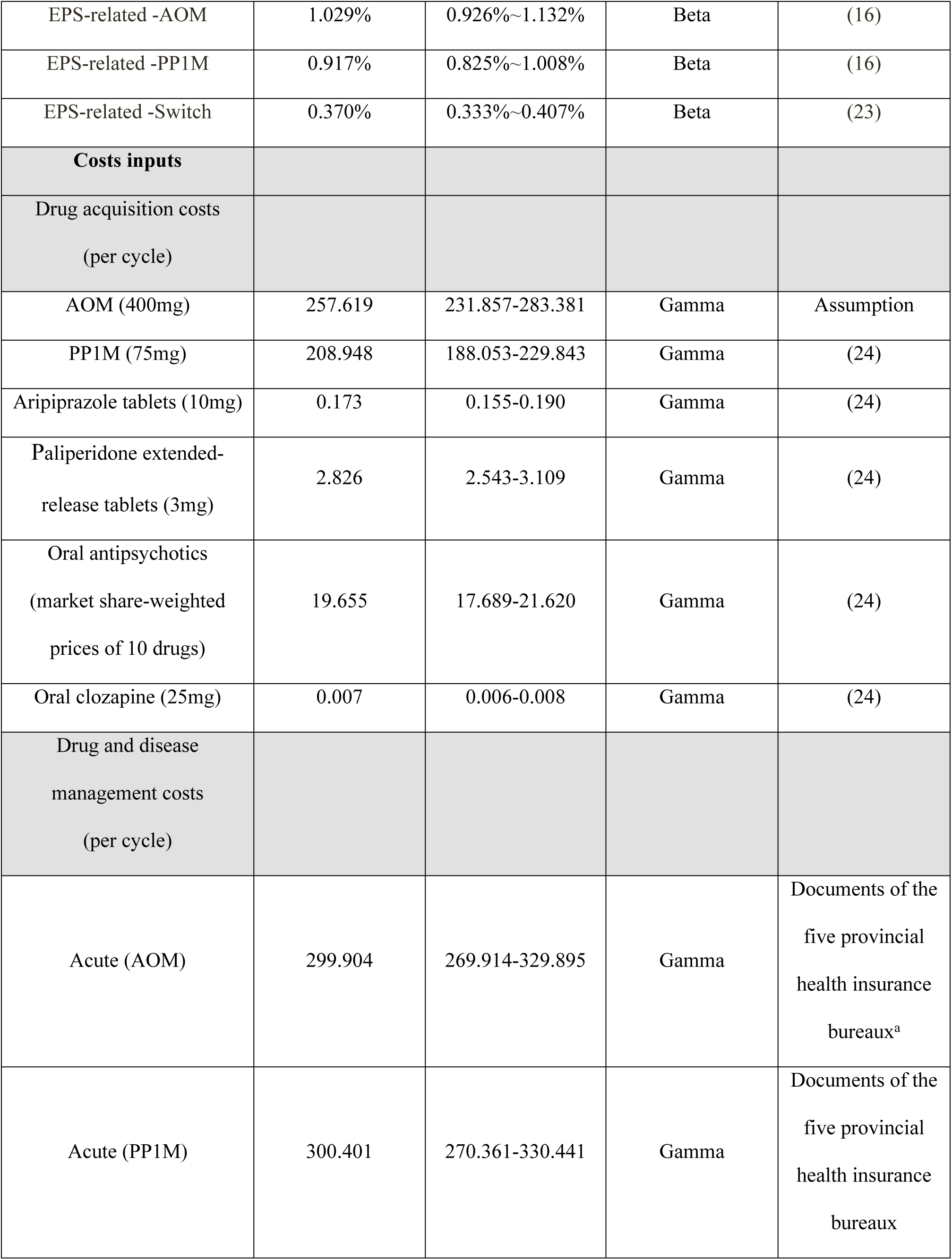
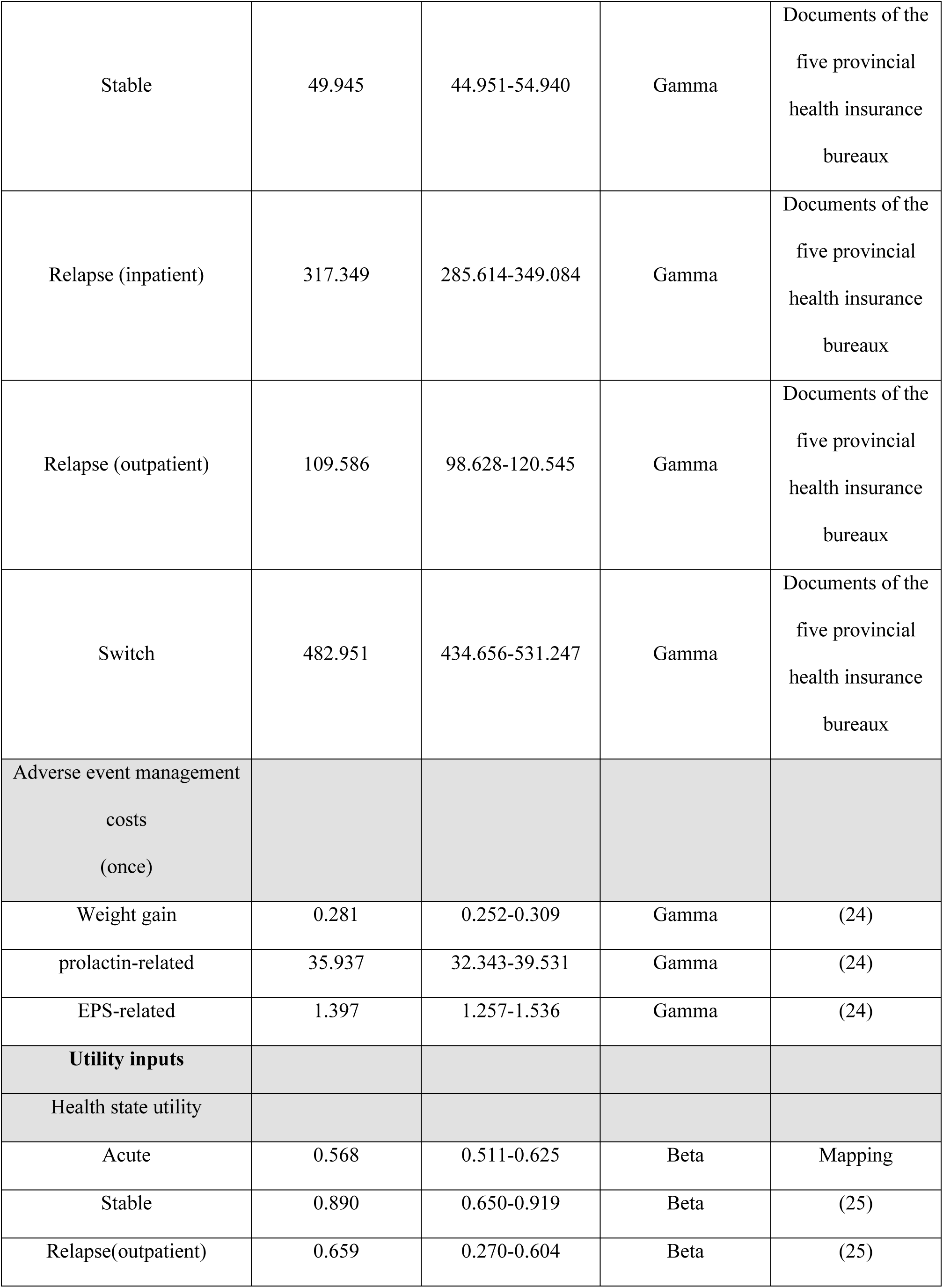
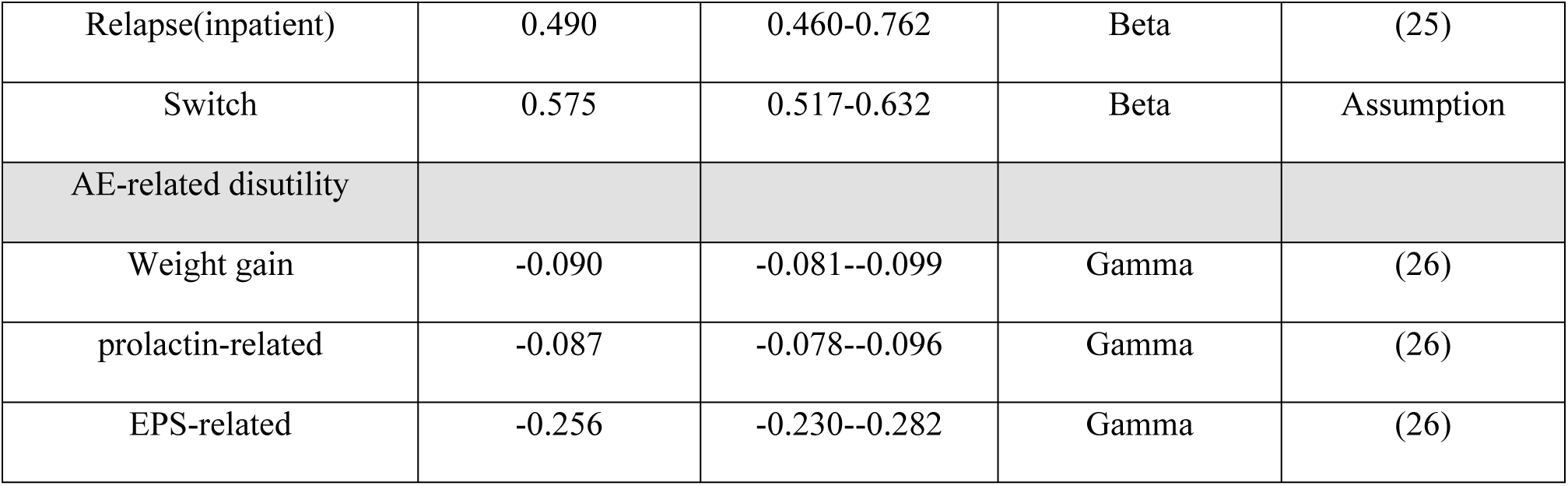
Model inputs.

| Parameters | Base-case value | Range | Distribution | References |
| --- | --- | --- | --- | --- |
| **Clinical inputs** |  |  |  |  |
| **AE rates in acute** |  |  |  |  |
| Weight gain-AOM | 23.200% | 20.880%~25.520% | Beta | MAIC |
| Weight gain-PP1M | 7.222% | 6.500%-7.944% | Beta | MAIC |
| Prolactin-related -AOM | 1.300% | 1.170%-1.430% | Beta | MAIC |
| Prolactin-related -PP1M | 1.385% | 1.247%-1.524% | Beta | MAIC |
| EPS-related -AOM | 40.600% | 36.540%-44.660% | Beta | MAIC |
| EPS-related -PP1M | 11.037% | 9.933%-12.141% | Beta | MAIC |
| **AE rates in relapse** |  |  |  |  |
| Weight gain-AOM | 8.423% | 7.581%-9.265% | Beta | MAIC |
| Weight gain-PP1M | 2.468% | 2.221%-2.715% | Beta | MAIC |
| Prolactin-related -AOM | 0.435% | 0.392%-0.479% | Beta | MAIC |
| Prolactin-related -PP1M | 0.464% | 0.418%-0.510% | Beta | MAIC |
| EPS-related -AOM | 15.939% | 14.345%-17.533% | Beta | MAIC |
| EPS-related -PP1M | 3.823% | 3.441%-4.206% | Beta | MAIC |
| **AE rates in stable & switch** |  |  |  |  |
| Weight gain-AOM | 2.107% | 1.896%~2.318% | Beta | (16) |
| Weight gain-PP1M | 3.335% | 3.002%~3.669% | Beta | (16) |
| Weight gain-Switch | 3.477% | 3.129%~3.825% | Beta | (23) |
| Prolactin-related -AOM | 0.000% | 0.000%~0.000% | Beta | (16) |
| Prolactin-related -PP1M | 1.021% | 0.919%~1.123% | Beta | (16) |
| Prolactin-related-Switch | 0.450% | 0.405%~0.495% | Beta | (23) |
| EPS-related -AOM | 1.029% | 0.926%~1.132% | Beta | (16) |
| EPS-related -PP1M | 0.917% | 0.825%~1.008% | Beta | (16) |
| EPS-related -Switch | 0.370% | 0.333%~0.407% | Beta | (23) |
| **Costs inputs** |  |  |  |  |
| Drug acquisition costs<br>(per cycle) |  |  |  |  |
| AOM (400mg) | 257.619 | 231.857-283.381 | Gamma | Assumption |
| PP1M (75mg) | 208.948 | 188.053-229.843 | Gamma | (24) |
| Aripiprazole tablets (10mg) | 0.173 | 0.155-0.190 | Gamma | (24) |
| Paliperidone extended-<br>release tablets (3mg) | 2.826 | 2.543-3.109 | Gamma | (24) |
| Oral antipsychotics<br>(market share-weighted<br>prices of 10 drugs) | 19.655 | 17.689-21.620 | Gamma | (24) |
| Oral clozapine (25mg) | 0.007 | 0.006-0.008 | Gamma | (24) |
| Drug and disease<br>management costs<br>(per cycle) |  |  |  |  |
| Acute (AOM) | 299.904 | 269.914-329.895 | Gamma | Documents of the<br>five provincial<br>health insurance<br>bureaux<sup>a</sup> |
| Acute (PP1M) | 300.401 | 270.361-330.441 | Gamma | Documents of the<br>five provincial<br>health insurance<br>bureaux |
| Stable | 49.945 | 44.951-54.940 | Gamma | Documents of the five provincial health insurance bureaux |
| Relapse (inpatient) | 317.349 | 285.614-349.084 | Gamma | Documents of the five provincial health insurance bureaux |
| Relapse (outpatient) | 109.586 | 98.628-120.545 | Gamma | Documents of the five provincial health insurance bureaux |
| Switch | 482.951 | 434.656-531.247 | Gamma | Documents of the five provincial health insurance bureaux |
| Adverse event management costs (once) |  |  |  |  |
| Weight gain | 0.281 | 0.252-0.309 | Gamma | (24) |
| prolactin-related | 35.937 | 32.343-39.531 | Gamma | (24) |
| EPS-related | 1.397 | 1.257-1.536 | Gamma | (24) |
| **Utility inputs** |  |  |  |  |
| Health state utility |  |  |  |  |
| Acute | 0.568 | 0.511-0.625 | Beta | Mapping |
| Stable | 0.890 | 0.650-0.919 | Beta | (25) |
| Relapse(outpatient) | 0.659 | 0.270-0.604 | Beta | (25) |
| Relapse(inpatient) | 0.490 | 0.460-0.762 | Beta | (25) |
| Switch | 0.575 | 0.517-0.632 | Beta | Assumption |
| AE-related disutility |  |  |  |  |
| Weight gain | -0.090 | -0.081--0.099 | Gamma | (26) |
| prolactin-related | -0.087 | -0.078--0.096 | Gamma | (26) |
| EPS-related | -0.256 | -0.230--0.282 | Gamma | (26) |

### Costs inputs

Direct medical costs encompass three primary components: drug acquisition costs, drug and disease management costs, and adverse event management costs. A summary of these costs is presented in Table 2.

The latest drug prices for each state treatment modality were obtained from China Pharmaceutical Information Database(24), with the exception of AOM, for which a simulated price was used. Drug and disease management costs were derived from publicly available data and expert research. The frequency of medical resource utilization by patients in different health states was determined based on psychiatrists’ opinions, including psychiatric clinical assessments, the number of psychiatric scale evaluations, hospitalization days, blood and urine tests, and electroencephalograms. (See appendix 5 in the S1 File for details of the costs.) The unit costs of these medical resources were derived from the average unit prices of medical projects published by the governments of five provinces in China.

Adverse events were managed in accordance with the literature.(27) Weight gain was addressed with metformin hydrochloride tablets, prolactin-related adverse events with bromocriptine mesylate tablets, and EPS-related adverse events with phenazopyridine hydrochloride tablets. The treatment duration for each was set at 14 days per cycle, based on expert research.

### Utility inputs

Utility and disutility inputs were shown in Table 2. Currently, there is a lack of schizophrenia utility studies specific to the Chinese population. To utilize the acute state population data from the AOM phase III trial, we employed the mapping method to estimate the acute state utilities. This method involves converting non-utility scale scores to utility scale scores using regression equations.(28) The mapping formula (Formula 2) was derived from a study based on an Asian population, with 59.8 % being Chinese, which mapped the PANSS score onto the five-level EuroQol five-dimensional (EQ-5D-5L) utilities. (29)

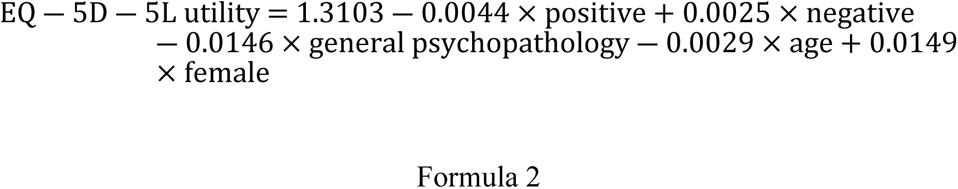

positive: positive subscales of PANSS, negative: positive subscales of PANSS, general psychopathology: subscales of PANSS

According to expert opinion, the utilities for the switch state are considered to be the average of the utilities for relapse outpatient and relapse inpatient states. This averaging approach was also employed in the absence of specific utilities for an 8-state Markov model study of schizophrenia.(18)

### Model Validation

We validated and refined the study design using the health economic quality evaluation tool CHEERS.(30) (See appendix 8 in the S1 File for details of CHEERS Checklist) During model construction, a targeted literature review of existing pharmacoeconomic studies of LAI for schizophrenia was conducted to ensure that the design of this study was generally consistent with previous studies. For internal validation, the discount rate for costs and utilities was set at 0 % to ensure that the undiscounted and discounted quality-adjusted life years (QALYs), as well as cost results, were consistent. For external validation, online and offline expert consultations were organized, employing semi-structured questionnaires and face-to-face interviews to gather opinions from psychiatric clinicians. This process aimed to ensure that the health state setting, simulation horizon, and treatment modalities for each state closely reflected clinical reality.

### Sensitivity analyses

A one-way sensitivity analysis was conducted to assess the impact of individual parameter changes on the base-case results. This analysis involved setting upper and lower limits on the model parameters, allowing each parameter to vary individually within 95 % confidence intervals reported in the literature or by ±10 % of the base-case values (when confidence intervals were not available). The annual discount rate was varied between 3 % and 8 %. Parameters varied included costs, utilities, and the incidence of adverse events, totaling 45 parameters. The results were depicted in the form of a tornado diagram, with parameters ranked in descending order based on the magnitude of their impact on net monetary benefits (NMB).

A probability sensitivity analysis was conducted using 1000 Monte Carlo simulations. The parameter distributions were specified with reference to ‘Cost-Effectiveness Modelling for Health Technology Assessment’.(17) Costs and disutility parameters were assumed to follow a Gamma distribution, while utilities and the incidence of adverse events parameters were assumed to follow a Beta distribution. The results were presented in the form of cost-effectiveness acceptability curves (CEACs) and incremental cost-effectiveness scatter plots.

### Scenario analyses

A systematic review and meta-analysis of life expectancy in patients with schizophrenia revealed that the life expectancy of Asian patients was 60.2 years (95 % CI: 56.6-63.8).(31) In contrast, the baseline mean age of patients with AOM and PP1M after MAIC was 31.5 years. Consequently, scenario analyses adjusted the simulation horizon of the study to a maximum of 30 years. The results were presented as line graphs.

## Result

### Base-case analysis

Table 3 presents the results of the base-case analysis. Over a 10-year horizon, the incremental costs of AOM compared to PP1M treatment were US$1,926.373, with an incremental QALY of 0.306 and an ICER of US$6,285.303/QALY. This ICER is below the WTP threshold of US$12,538.306. In the acute state, the total costs of AOM are lower than those of PP1M, and both have a QALY of 0.12, making AOM the dominant strategy in short-term 12-week treatments. In other health states, AOM has a larger population in the stable state, and cumulative treatments generate more QALYs than PP1M. (See appendix 7 in the S1 File for details of cost and utility components.)

**Table 3.** Base-case analysis.

|  | **AOM** | **PP1M** |
| --- | --- | --- |
| **Costs outcomes (US\$)** | | |
| Drug acquisition costs | 25,464.029 | 19,146.105 |
| Drug and disease management costs | 11,684.268 | 15,549.140 |
| Adverse event management costs | 6.853 | 36.218 |
| Total costs(discounted) | 36,494.821 | 34,568.448 |
| **Health outcomes** |  |  |
| Treatment increased QALYs | 7.552 | 7.235 |
| AE decreased QALYs | -0.032 | -0.027 |
| Total QALYs (discounted) | 7.384 | 7.077 |
| **Incremental results** |  |  |
| Incremental costs (US\$) | 1,926.373 | - |
| Incremental QALYs | 0.306 | - |
| ICER (US\$/QALYs) | 6285.303 | - |

Although the loss of QALYs due to adverse events is slightly greater for AOM than for PP1M, this represents a cognitive reversal of the 0.306 incremental QALYs associated with AOM. Nonetheless, the adverse event management costs and the loss of QALYs have a minimal impact on the overall outcome, as confirmed by previous pharmacoeconomic studies.(32) This study did not include adverse event treatment costs in the model. The incremental QALYs in this study were mainly due to the superior performance of AOM in terms of responder rate, and AOM is also cost-effective in the long term.

### Sensitivity analyses

The results of the one-way sensitivity analysis are depicted in Fig 2, which only shows the top 10 parameters influencing the results due to the large number of parameters considered. The parameter exerting the greatest influence on Net Monetary Benefits (NMB) is the cost of AOM drug acquisition, followed by the cost of PP1M drug acquisition.

**Figure 2.**
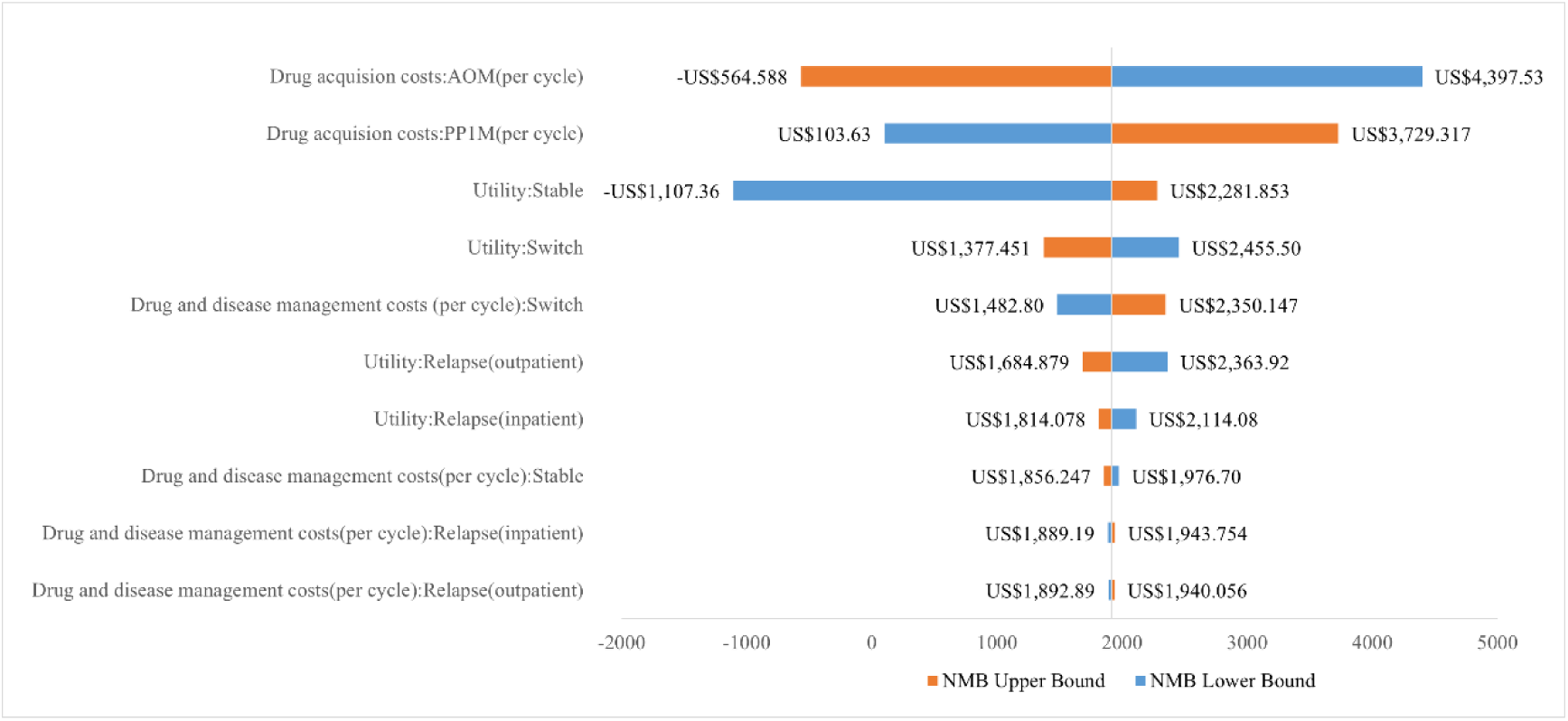
One-way sensitivity analysis.

The results of the probability sensitivity analysis, conducted after 1000 Monte Carlo simulations, are presented in Figs 3 and 4. According to the cost-effectiveness acceptability curve, the probability that AOM is more economical under the WTP threshold in China for 2023 is 92.6 %, indicating that the results of the base-case analysis are robust.

**Figure 3.**
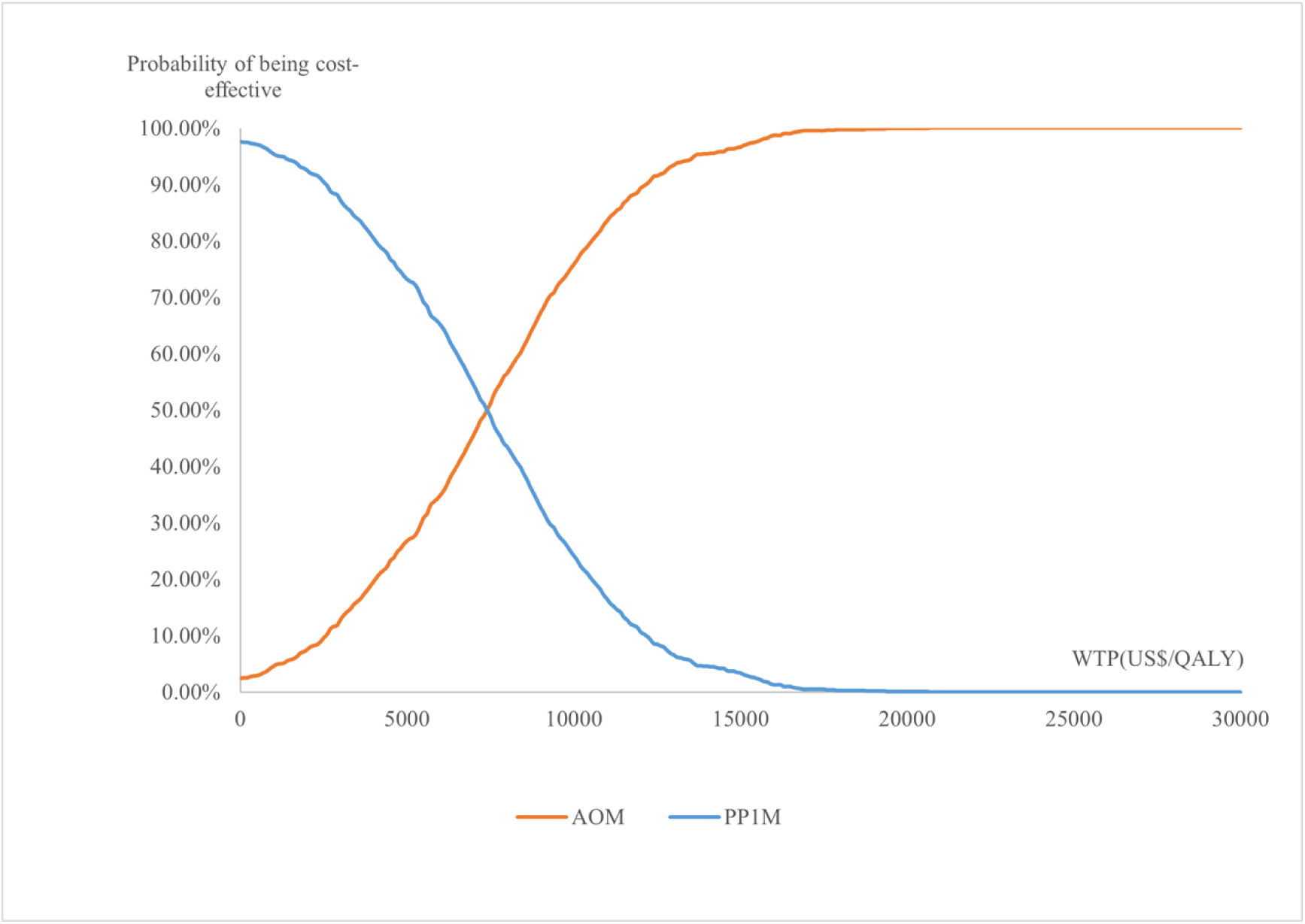
Cost-effectiveness acceptability curve.

**Figure 4.**
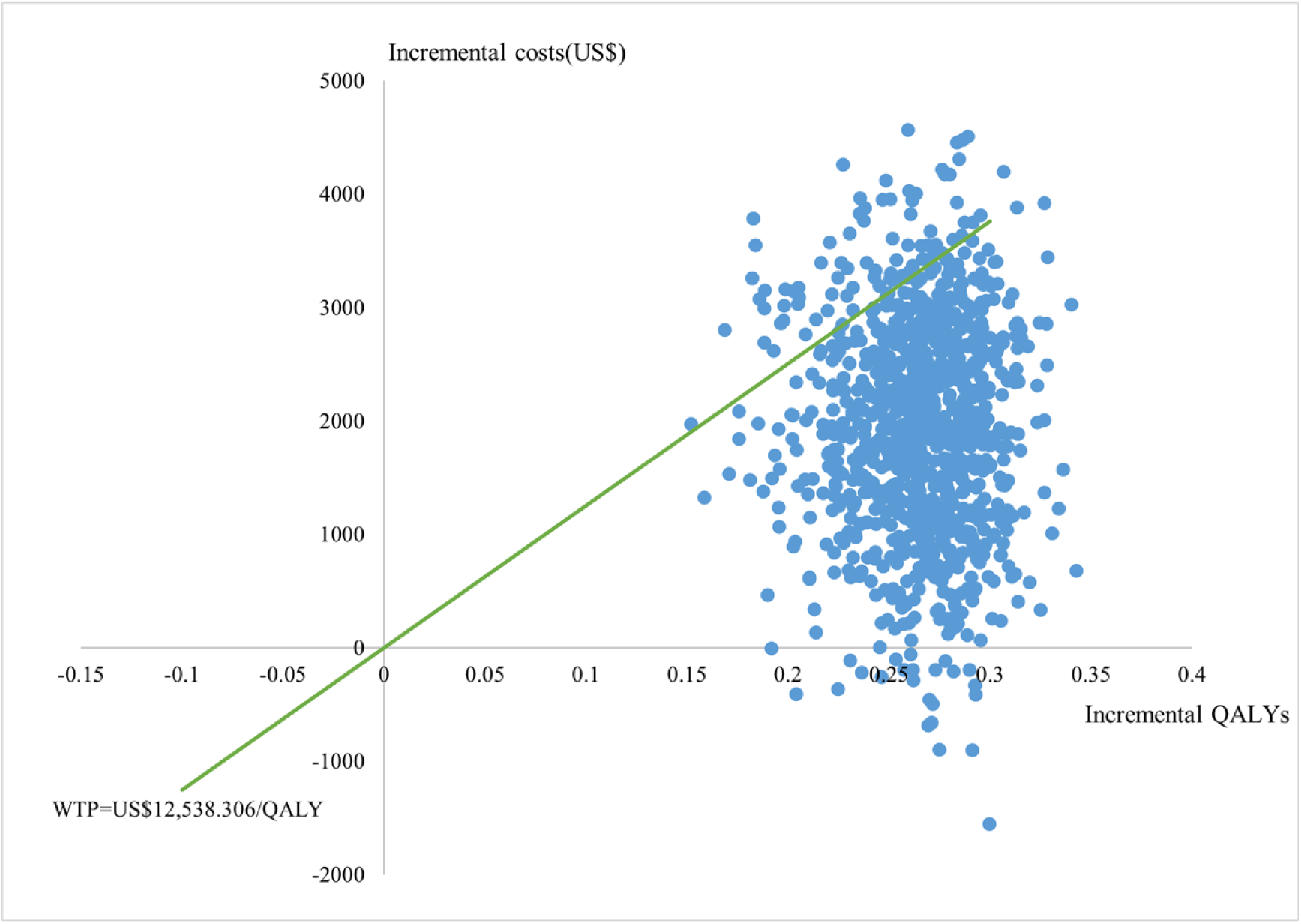
Incremental cost-effectiveness scatter plot.

### Scenario analyses

Over a 30-year horizon, the incremental costs of AOM compared to PP1M treatment were US$4,522.573, with an incremental QALY of 0.789 and an ICER of US$5,733.204/QALY. This ICER is below the WTP. The results of the scenario analysis were shown in Fig 5. The ICER began to fall below the WTP after one month of treatment and remained beneath this threshold, thereby demonstrating the economic advantage of the long-term use of AOM.

**Figure 5.**
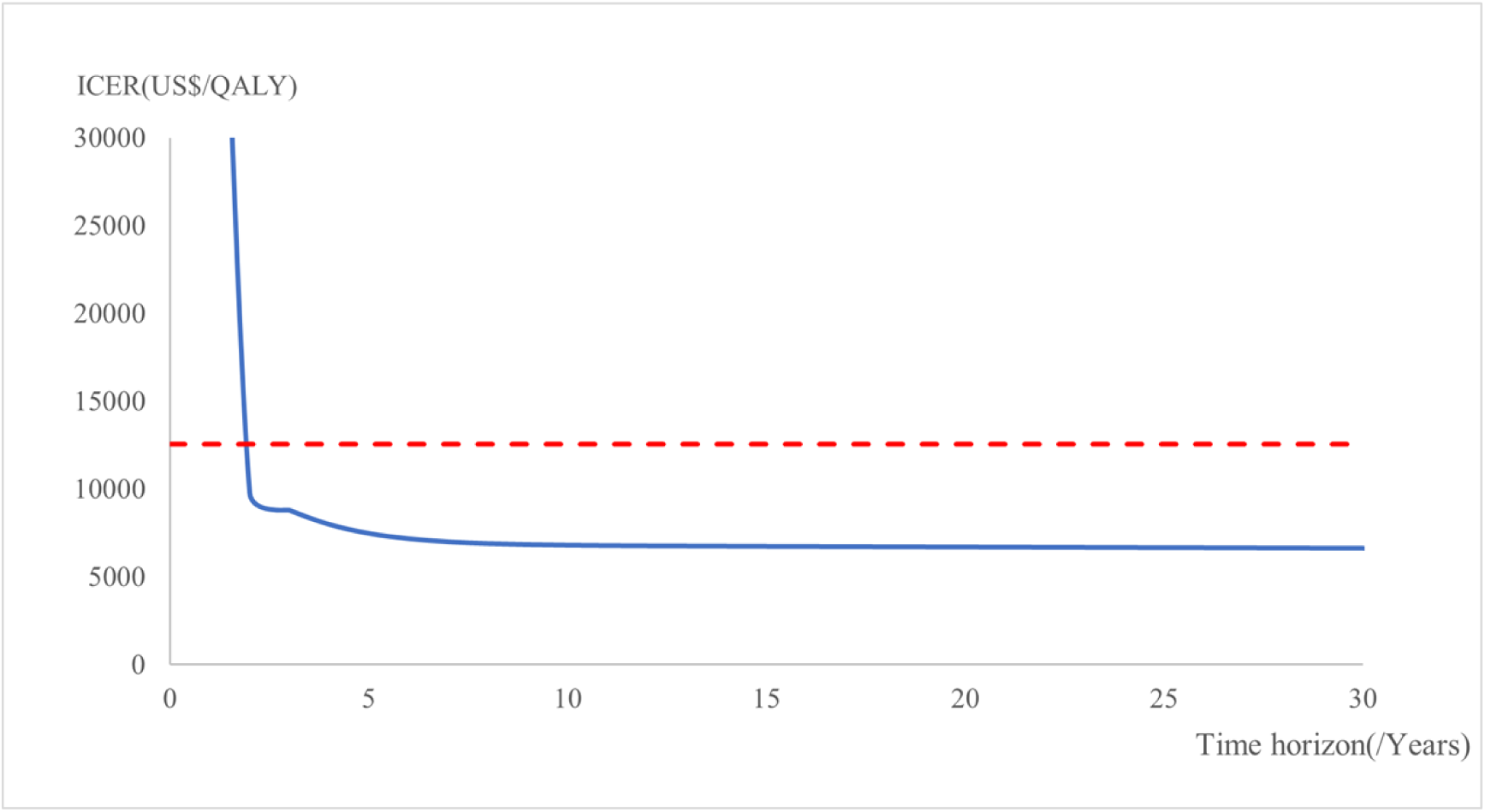
ICER changes with time horizon.

## Discussion

In this study, we constructed a 5-state Markov model to evaluate the cost-utility of AOM versus PP1M for the treatment of schizophrenia in Chinese adults from the perspective of the Chinese healthcare system. We reviewed the pharmacoeconomic evidence for these two long-acting injectables to ensure that the conclusions of our study were consistent with those of previous studies. Leslie Citrome et al (33) in the United States constructed a 1-year decision tree model for cost-effectiveness analysis. Clinical inputs were obtained from the placebo-controlled trials of AOM and PP1M.(34, 35) The relapse rate in our study was also obtained from these two trials, which are the only available data currently. Their results showed that AOM was dominated at real-world and maximum doses, and the ICER was below the WTP at both clinical trial doses and prescription doses. Notably, Leslie Citrome et al. used the relapse rate data from the original trial directly as model inputs, whereas our study conducted a meta-analysis on his basis. Christophe Sapin et al (36) in the United States constructed a 6-month covariance model for cost-effectiveness analysis. Clinical inputs were obtained from the QUALIFY study. The result demonstrated that AOM was dominant over PP1M in improving QLS and CGI-S scores. A cost-effectiveness analyses in France also based on the QUALIFY study reached the same conclusions. (37)

The economic evidence pertaining to AOM and PP1M is relatively limited. The existing studies were conducted in developed countries and focused on short-term cost-effectiveness analysis with a time horizon of 6 months to 1 year. The primary clinical inputs were derived from the QUALIFY study, which recruited participants from 10 developed countries, and the effect outcomes CGI scores and QLS scores were not conducive to the interpretation of pharmacoeconomic results (costs per subscale score improvement). These evidences are less applicable in developing countries.

Our study represents the first long-term cost-utility analysis conducted within the context of a developing country’s healthcare system, with a study horizon of up to 30 years for AOM and PP1M. Clinical inputs for the acute state were derived from the Chinese population for both the AOM and PP1M trials, and acute state utilities were derived from a mapping study that also included a Chinese population. Although the clinical inputs and utilities in other health states were sourced from foreign populations, we have used the available Chinese population data for model inputs as much as possible to make the model more relevant to the Chinese reality. We used utility analysis to make the results more understandable to policy makers (costs per healthy year of life).

However, this study also has the following limitations: first, the use of model parameters was limited by the scarcity of data from head-to-head clinical trials of AOM and PP1M and the short study time frame. In this study, we chose the AOM phase III positive double-blind controlled trial and the PP1M phase IV single-arm open-label trial for MAIC to obtain acute state parameters, and there were differences in the design of the clinical trials themselves, and there was some bias between the results after MAIC and the reality. Most clinical studies have observed fewer relapse events in patients using LAI during the follow-up period, and there is a lack of data for the relapse population. This study hypothesized that the efficacy and safety of the relapse state would be consistent with those of the acute state, this hypothesis requires validation with data from a broader relapse population.

Second, in the meta-analysis of stable state relapse rate, we targeted the specifications of AOM and PP1M as 1-month injections. Only two clinical trials met the requirements after rigorous screening according to the inclusion and exclusion criteria and bias evaluation, which is a weak level of evidence. The current large scale meta-analysis in the field of psychiatric disorders often combined bipolar disorder other than schizophrenia, depression, and other disorders, and included LAI and oral dosage forms and different specifications of drugs. These have limited reference value for the present study.

Third, the model’s utilities except for the acute state were derived from extra-domain studies with non-uniform sources of utilities, and despite we did sensitivity analyses for this, variability between populations may still lead to bias. There are fewer acute state utility studies, and the acute state utilities used in published pharmacoeconomic studies were not clearly sourced, so we did not refer to their value for a scenario analysis of different utility sources.

Fourth, both AOM and PP1M are LAI, so the difference in adherence between the two drugs was not considered in this study, however, for the clinical reality of long-term treatment, patients still discontinue their medication. The model simplifies this aspect, potentially leading to an overestimation of total costs.

We have tried our best to narrow these gaps through reasonable hypotheses and study design, but it still leads to limited extrapolation of the conclusions of this study. We hope that more long-term prospective trials or retrospective data will be available in the future to demonstrate the clinical differences between the two drugs, and we also hope that there will be local Chinese studies on the health-related quality of life of schizophrenia patients. Although LAI for schizophrenia have been put into clinical use in China for years, the clinical utilization rate is only 0.66 %, much lower than the Asian average of 17.9 %.(38) We hope that this study will support the AOM health care access decision, promote the widespread clinical use of LAI, improve mental health management in China, and provide reference for health care decisions in developing countries.

## Conclusion

In the Chinese healthcare system perspective, the ICER of AOM is below the willingness-to-pay threshold for both 10 and 30 years of treatment, and is economically viable in the long term compared to PP1M.

## Data Availability

All relevant data are within the manuscript and its Supporting Information files.

No

## Contributors

An is first author. Ding and Li are the leader of the study, provided critical guidance. An, Fang and Zhang collected the data related to schizophrenia, designed the study. An built the Markov model, analyzed the data and written paper. All authors have read and approved the final manuscript. Ding and Li are the study guarantors.

## Data sharing

Data available in manuscript and supplementary materials.

## Supporting information

S1 File. This is the S1 File: Cost-utility of aripiprazole once-monthly versus paliperidone palmitate once-monthly injectable for schizophrenia in China Supporting information.

